# Parental inflammatory bowel disease and autism in the offspring: Triangulating the evidence using four complementary study designs

**DOI:** 10.1101/2021.06.09.21258393

**Authors:** Aws Sadik, Christina Dardani, Panagiota Pagoni, Alexandra Havdahl, Evie Stergiakouli, Jakob Grove, Golam M. Khandaker, Sarah A. Sullivan, Stan Zammit, Hannah J. Jones, George Davey Smith, Christina Dalman, Håkan Karlsson, Renee M. Gardner, Dheeraj Rai

## Abstract

**Importance:** Evidence linking parental diagnoses of inflammatory bowel disease (IBD) with offspring autism is inconclusive.

**Objective:** To investigate associations between parental diagnoses of IBD and offspring autism and elucidate their underlying aetiology by conducting four complementary studies.

**Design, Setting and Participants:** (1) Nationwide population-based cohort study using Swedish registers to examine associations between parental IBD diagnoses and autism diagnoses in offspring, (2) Linkage disequilibrium (LD)-score regression to estimate the genetic correlation between the phenotypes. (3) Polygenic risk score (PRS) analyses in the Avon Longitudinal Study of Parents and Children (ALSPAC) to investigate associations between maternal genetic liability to IBD and autism factor mean score in offspring. (4) Two-sample Mendelian randomization (MR) to assess bidirectional causal links between genetic liability to IBD and autism.

**Results:** Observational analyses provided evidence of an association between parental IBD diagnoses and offspring autism diagnosis in mutually adjusted models (maternal: OR= 1.32; 95% CI: 1.25 to 1.40; p<0.001; paternal: OR= 1.09; 95% CI: 1.02 to 1.17; p=0.012; n=2 324 227, 52.3% male). PRS analyses in ALSPAC indicated associations between maternal PRS for IBD subtypes and a measure of broad autism phenotype, autism factor mean score, in the offspring (UC: β_PRS_= 0.02; 95%CI: 0.003 to 0.05; p= 0.02; R^2^=0.06; Crohn’s: β_PRS_= 0.03; 95%CI: 0.01 to 0.05; p= 0.004; R^2^= 0.06; n= 7357, 50.3% male). MR analyses provided evidence of a potential causal effect of genetic liability for IBD, especially ulcerative colitis, on autism (OR_MR_= 1.03; 95%CI: 1.01 to 1.06). There was little evidence to suggest a causal effect of genetic liability to autism on risk of IBD, or a genetic correlation between the two conditions.

**Conclusions and relevance:** Triangulating evidence from a nationwide register-based cohort study, genetic correlation, polygenic risk score analyses and MR, we found evidence of a potentially causal link between parental, particularly maternal, diagnoses and genetic liability to IBD and offspring autism. Perinatal immune system dysregulation, micronutrient malabsorption and anaemia may be implicated.

## INTRODUCTION

Autism spectrum disorder (autism) is a long-term neurodevelopmental condition with a highly variable clinical manifestation^1^. A substantial proportion of individuals with autism diagnoses present with gastrointestinal symptoms^2,3^, leading to interest in potential associations with gastrointestinal conditions, such as inflammatory bowel disease (IBD). In particular, a link between parental IBD and offspring autism has been hypothesised. There is evidence suggesting that autism and IBD might share risk genes^4^, while characteristics of IBD such as immune system dysregulation, micronutrient malabsorption, and anaemia^5–7^ may be perinatal risk factors for autism^8–11^. However, observational studies have provided inconclusive evidence^12–15^, and the underlying aetiology of any associations is unclear. Elucidating the associations between parental IBD and offspring autism may offer mechanistic insights into the origins of autism.

We conducted four complementary studies (Figure 1) to investigate: (1) associations between parental diagnoses of IBD and offspring autism in a nationwide cohort in Sweden; (2) genetic correlation between IBD and autism using genome-wide association study (GWAS) summary statistics; (3) polygenic associations between maternal genetic liability to IBD and offspring autistic traits in a large UK birth cohort; and (4) potential causal effects of genetic liability to IBD on autism and the possibility of reverse causation using bidirectional two-sample Mendelian randomization (MR).

**Figure 1.**
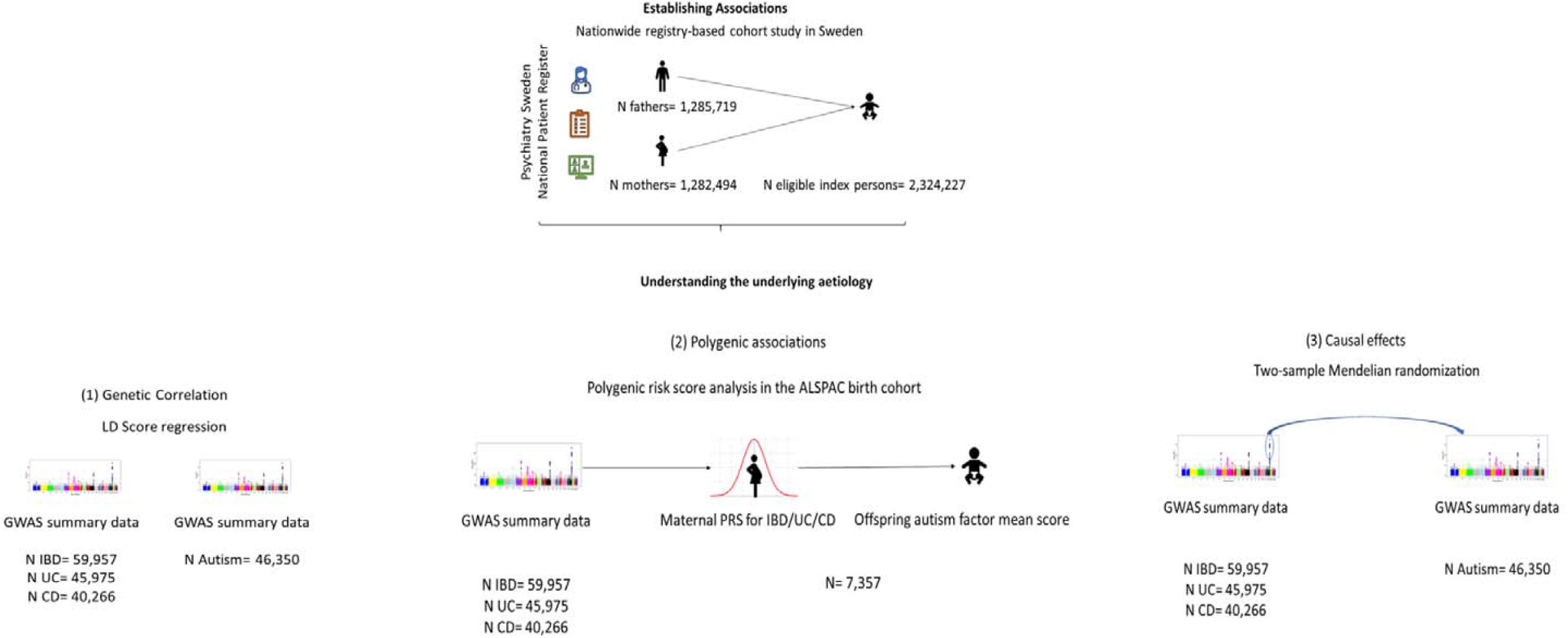
Summary of studies conducted in the present study, aiming at investigating the links between parental diagnoses of IBD and offspring autism and elucidating their underlying aetiology.

## METHODS

### Study 1: Investigating associations between parental diagnoses of IBD and offspring autism-Swedish cohort study

We used individual-level data from ‘Psychiatry Sweden’, a comprehensive national register linkage, to investigate whether parental IBD diagnosis is associated with offspring autism diagnosis.

All children born in Sweden from 1-January-1987 to 31-December-2010 (n= 2,837,045) were eligible index persons, with follow-up to 31-December-2016. Exclusion criteria were: children born outside Sweden (n=292,023), not registered in the Medical Birth Register (n=74,240), resident in Sweden for <5 years (n=23,495), multiple pregnancy (n=67,309), adopted (n=2,425), known genetic/metabolic causes of neurodevelopmental conditions (e.g. trisomies) (n=7,873) or incomplete parental records (n=45,453)^16^. The study population included 2,324,227 offspring born to 1,282,494 mothers and 1,285,719 fathers (Supplementary Figure S1). The Stockholm Regional Ethical Review Committee (DNR 2010/1185-31/5) approved the study.

Offspring autism was identified in the National Patient Register (NPR) using ICD-9 and ICD-10 codes (eMethods S1). Lifetime history of parental IBD, Crohn’s disease (Crohn’s) and ulcerative colitis (UC) were identified using ICD-9 and ICD-10 codes in the NPR (eMethods S1).

Using STATA/MP15, we estimated the odds ratios and 95% confidence intervals of the association of mother’s and father’s diagnosis of IBD (any IBD, Crohn’s, or UC) with offspring autism using generalised estimating logistic models with robust standard errors accounting for clustering of multiple children born to the same parents.

Model 1 was unadjusted. Model 2 was adjusted for parental age at delivery^17^, migrant status^18^, education level, family income quintile at birth^19^, parents’ history of psychiatric diagnosis prior to the birth of the child and offspring sex, birth year and birth order. Model 3 was additionally mutually adjusted for maternal and paternal IBD diagnoses to avoid bias from assortative mating^20^. As a sensitivity analysis, we restricted parental IBD diagnoses to those recorded prior to the birth of the index person and investigated associations with offspring autism. Additionally, we investigated associations between any parental IBD diagnoses and offspring autism with and without intellectual disabilities (ID) separately, since these groups may have distinct genetic and environmental risk factors^21–24^ and outcomes^25,26^.

### Study 2: Investigating genetic correlations-LD-Score regression

We used LD-score regression to estimate the genetic correlation between genetic liability to autism and IBD, Crohn’s and UC.

LD-score regression allows the estimation of the genetic correlation between polygenic traits using GWAS summary statistics by capitalising on patterns of linkage disequilibrium among common genetic variants^27^. We used the latest available GWAS summary data on autism (N_cases_= 18,381; N_controls_= 27,969)^28^, IBD (N_cases_= 25,042; N_controls_= 34,915)^29^, Crohn’s (N_cases_=12,194; N_controls_= 28,072)^29^ and UC (N_cases_= 12,366; N_controls_= 33,609)^29^. Detailed information on study samples and case definition can be found in the original publications.

We followed the suggested protocol for LD-score regression analyses (https://github.com/bulik/ldsc/wiki). Using the LDSC (LD Score) v1.0.1 software in Python, we estimated genetic correlations using pre-computed LD scores from the 1000 Genomes project European data^30^ (from: https://data.broadinstitute.org/alkesgroup/LDSCORE/eur_wld_chr.tar.bz) with an unconstrained intercept term to account for any sample overlap, and population stratification.

### Study 3: Investigating associations between genetic liability to IBD and childhood broad autism phenotype-Polygenic Risk Score analysis in mothers and children of the ALSPAC cohort

#### Discovery Sample

Common genetic variants, corresponding alleles, effect sizes and p-values were extracted in order to calculate polygenic risk scores (PRSs), from the GWAS summary data of IBD^29^, UC^29^ and Crohn’s^29^ described above.

#### Target Sample

ALSPAC is a UK prospective birth cohort study based in Bristol and surrounding areas, which recruited pregnant women with expected delivery dates from 1 April 1991 to 31 December 1992; 14,541 women were initially enrolled, with 14,062 children born, and 13,988 children alive at 1 year of age. Detailed information on the cohort is available elsewhere^31,32^. A fully searchable study data dictionary is available at : http://www.bristol.ac.uk/alspac/researchers/our-data/. Ethical approval for the study was obtained from the ALSPAC Ethics and Law Committee and the Local Research Ethics Committees.

##### Genetic data

10,015 ALSPAC mothers were genotyped on the Illumina Human660W-quad genome-wide single nucleotide polymorphism (SNP) genotyping platform, and 9,912 ALSPAC children were genotyped on the Illumina HumanHap550-quad. After standard quality control (eMethods S2) and excluding participants who had withdrawn consent, genetic data were available for 7,921 mothers and 7,977 children of European ancestry. Consent for biological samples has been collected in accordance with the Human Tissue Act (2004).

##### Broad autism phenotype-autism factor mean score

We used a measure of the broad autism phenotype previously estimated in ALSPAC as the mean score of 7 factors derived from a factor analysis of 93 measures related to autism in ALSPAC^33^. The measure was available in 13,128 children and strongly predictive of the autism diagnosis measured independently via school records, record linkage and parental reports^33^. Other autism trait measures or diagnoses were not used as there were fewer genotyped mothers and children with these measures.

##### Calculation of Polygenic Risk Scores in ALSPAC and statistical analysis

PRS were calculated using PLINK version 1.9, applying the method described by the Psychiatric Genomics Consortium (PGC)^34^. SNPs with mismatching alleles between the discovery and target dataset were removed. The MHC region was removed (25 Mb – 34 Mb), except for one SNP representing the strongest signal within the region. Using ALSPAC data as reference panel, SNPs were clumped with an r^2^ of 0.25 and a physical distance threshold of 500 kB. The optimal p-value threshold for PRS is dependent on discovery and target sample sizes, as well as SNP inclusion parameters (e.g., r^2^)^35,36^. For this reason, we calculated PRS for each participant across 13 p-value thresholds (5e-8 to 0.5), standardised by subtracting the mean and dividing by the standard deviation. We defined PRS corresponding to p-value threshold 0.05 as our primary exposure, based on a previous ALSPAC study^37^. This threshold has been found to have sufficient predictive ability for IBD and its subtypes^38^. We could not directly assess the predictive power and optimal p-value threshold of our PRSs in our target sample as there were few UC (n=12) and Crohn’s cases (n=16).

After constructing PRS for IBD, UC and Crohn’s in mothers and children, we performed linear regressions using STATA/MP 15 to examine associations with the standardised autism factor mean score in childhood. Analyses were adjusted for child’s sex and the first 10 principal components of the ALSPAC genotype data to avoid population stratification bias^35^.

### Study 4: Investigating bidirectional causal links-Two-sample Mendelian randomisation

We performed two-sample Mendelian randomisation (MR) to assess bidirectional causal links between genetic liability to autism and IBD and its subtypes, and vice versa.

MR can be implemented as an instrumental variable approach, utilising common genetic variants as instruments for exposures of interest, allowing assessment of causal effects and their direction on outcomes. MR relies on the following assumptions : (i) there must be a robust association between the common genetic variants and the exposure, (ii) the variants should operate on the outcome entirely via the exposure, (iii) the variants should not be associated with any confounders of the associations between the exposure and the outcome^39^. In this study, we applied two-sample MR, in which the effect sizes and standard errors of the instruments for the exposure and the outcome were extracted from separate GWASs conducted in independent samples from the same underlying population^40^.

#### Genetic Instruments

Genetic instruments were extracted from the overlapping set of SNPs between the autism^28^, IBD^29^, UC^29^, and Crohn’s^29^ GWASs. This ensured that all selected genetic instruments would be present in the outcome GWAS.

GWAS summary data were restricted to a common set of SNPs and then clumped in PLINK 1.90 using the 1000Genomes^30^ phase 3 European ancestry reference panel, and an r2= 0.01, within a 10,000 kb window. Among the independent variants, instruments were defined using a genome-wide significance threshold of p≤5×10^−08^. The only exception was autism, as only two independent and genome-wide significant variants were identified. We therefore relaxed the p-value threshold to 5×10^−07^ to improve statistical power, as used previously ^41^. Supplementary Figure S1 illustrates the process of instrument definition, and supplementary table S1 contains information on the genetic instruments used.

#### Harmonisation

We harmonised the alleles of the outcome on the exposure, to ensure SNP-exposure and SNP-outcome effects correspond to the same allele. Variants identified as palindromic were removed, as the effect allele frequencies in the IBD, UC, and Crohn’s GWASs were not provided. Supplementary tables S2 and S3 contain details of the harmonised datasets.

#### Inverse Variance Weighted MR

The primary MR analysis was the Inverse Variance Weighted (IVW) method which provides an overall causal effect estimate of the exposure on the outcome, estimated as a meta-analysis of the ratios of the SNP-outcome effect to the SNP-exposure effect weighted by each SNP’s relative precision^42^.

#### Sensitivity Analyses to test robustness of causal effect estimates

We assessed the strength of the instruments by estimating the mean F statistic. As a rule of thumb, the IVW is unlikely to suffer from weak instrument bias if mean F>10^44^.

We assessed the consistency of the IVW causal effect estimates using sensitivity analyses, including: MR Egger regression^42^, Weighted Median^45^ and Weighted Mode^46^ (eMethods S3).

#### Sensitivity Analyses to test the consistency of the causal effect estimates in autism without intellectual disabilities (ID)

The autism GWAS used in our primary analyses included a proportion of autism cases with ID^28^. We tested the consistency of the causal effect estimates using GWAS summary data on a sub-sample of the iPSYCH cohort^47^ excluding all intellectual disability cases (N_cases_= 11,203; N_controls_= 22,555). Supplementary figure S2 visualises the process of instrument definition, and supplementary tables S4, S5 and S6 contain details on the instruments used and the harmonised datasets.

Two-sample MR analyses were performed using the TwoSampleMR R package^43^ in R version 3.5.1.

## RESULTS

### Study 1: Associations between parental IBD diagnoses and offspring autism

Cohort characteristics by exposure to parental IBD diagnoses are presented in supplementary tables S7 and S8.

Maternal IBD diagnosis was associated with offspring autism in crude and adjusted models (Any IBD diagnosis: OR_MODEL3_= 1.32; 95% CIs: 1.25 to 1.40; Table 1). Similar results were observed in analyses of maternal UC and Crohn’s diagnoses and offspring autism (Table 1), and in analyses restricted to maternal IBD diagnoses prior to the index person’s birth (Any IBD diagnosis: OR_MODEL3_= 1.20; 95% CIs: 1.09 to 1.32; Supplementary Table S9). The paternal IBD associations with autism were weaker (OR_MODEL3_= 1.09; 95% CIs 1.02 to 1.17) than the maternal associations (Table 1). Point estimates for associations of parental IBD diagnoses to autism without ID were higher than those for autism with ID, although confidence intervals overlapped (Supplementary Table S10).

**Table 1.** Associations between maternal or paternal diagnosis for any inflammatory bowel disease (IBD), ulcerative colitis, Crohn’s disease and offspring diagnosis of autism.

| Exposure | Maternal diagnoses |  |  |  |  |  | Paternal diagnoses |  |  |  |  |  |
| --- | --- | --- | --- | --- | --- | --- | --- | --- | --- | --- | --- | --- |
|  | Model1<sup>a</sup><br>OR<br>(95% CIs) | P | Model2<sup>b</sup><br>OR<br>(95% CIs) | P | Model3<sup>c</sup><br>OR<br>(95% CIs) | P | Model1<sup>a</sup><br>OR<br>(95% CIs) | P | Model2<sup>b</sup><br>OR<br>(95% CIs) | P | Model3<sup>c</sup><br>OR<br>(95% CIs) | P |
| **Any IBD** | 1.39<br>(1.31,1.47) | <0.001 | 1.32<br>(1.24,1.40) | <0.001 | 1.32<br>(1.25,1.40) | <0.001 | 1.14<br>(1.06,1.22) | <0.001 | 1.11<br>(1.03,1.18) | 0.004 | 1.09<br>(1.02,1.17) | 0.012 |
| **Crohn's Disease** | 1.23<br>(1.09,1.40) | 0.001 | 1.19<br>(1.05,1.35) | 0.006 | 1.20<br>(1.06,1.36) | 0.004 | 1.18<br>(1.04,1.35) | 0.013 | 1.16<br>(1.02,1.33) | 0.023 | 1.16<br>(1.01,1.32) | 0.031 |
| **Ulcerative Colitis** | 1.24<br>(1.12,1.38) | <0.001 | 1.22<br>(1.10,1.35) | <0.001 | 1.22<br>(1.10,1.36) | 0.001 | 0.99<br>(0.88,1.10) | 0.806 | 0.98<br>(0.87,1.09) | 0.662 | 0.97<br>(0.86,1.08) | 0.575 |

<sup>a</sup> Crude models.
<sup>b</sup> Models adjusted for child's sex, year of birth, birth order, maternal/paternal age, migrant status, education level, family income and parental psychiatric history.
<sup>c</sup> Mutually adjusted models for maternal/paternal IBD diagnoses, child's sex, year of birth, birth order, maternal/paternal age, migrant status, education level, family income and parental psychiatric history.
OR= odds ratio; CI = confidence interval

### Study 2: Genetic correlation between IBD and autism

We found little evidence of a genetic correlation between genetic liability to autism and IBD, UC, or Crohn’s (Table 2). Heritability scores, chi-squares and intercepts satisfied the conditions to provide reliable LD-score regression estimates^27,48^ (Supplementary Table S11).

**Table 2.** LD-score regression coefficients (r_g_), 95% confidence intervals (95% CIs) and p-values for the analyses investigating the genetic correlation between genetic liability to autism, Inflammatory Bowel Disease (IBD), ulcerative colitis and Crohn’s disease.

| Trait 1 | Trait 2 | $r_g$ (95% CIs) | P |
| --- | --- | --- | --- |
| Autism | IBD | -0.0615 (-0.15, 0.02) | 0.158 |
| Autism | Ulcerative colitis | -0.0656 (-0.17, 0.04) | 0.2064 |
| Autism | Crohn's disease | -0.0403 (-0.13, 0.05) | 0.3551 |

### Study 3: Associations between polygenic risk for IBD, UC, Crohn’s and broad autism phenotype in ALSPAC

The study sample derivation and characteristics can be found in Supplementary Figure S4.

#### Maternal polygenic risk for IBD, UC, Crohn’s and offspring broad autism phenotype

Maternal polygenic risk for UC and Crohn’s was associated with a higher autism factor mean score in the offspring (UC: β_PRS_= 0.02; 95%CIs: 0.003 to 0.05; p= 0.02; Crohn’s: β_PRS_= 0.03; 95%CIs: 0.01 to 0.05; p= 0.004). Similar results were found across other p-value thresholds (0.5-0.05). The effect size of the association between maternal polygenic risk for IBD and autism factor mean score, was comparable to that of UC and Crohn’s, although confidence intervals crossed the null (β_PRS_= 0.02; 95%CIs: -0.003 to 0.04; p= 0.09; R^2^= 0.06; Table 3, Supplementary Figure S5, Supplementary Table S12).

**Table 3.** Associations between child and maternal PRS for inflammatory bowel disease (IBD), ulcerative colitis, Crohn’s disease at p-value threshold 0.05, and autism factor mean score in the children of the ALSPAC birth cohort.

|  | IBD PRS |  |  |  | Ulcerative colitis PRS |  |  |  | Crohn's disease PRS |  |  |  |
| --- | --- | --- | --- | --- | --- | --- | --- | --- | --- | --- | --- | --- |
|  | Mother<br>N= 7,357 |  | Child<br>N= 7,506 |  | Mother<br>N= 7,357 |  | Child<br>N= 7,506 |  | Mother<br>N= 7,357 |  | Child<br>N= 7,506 |  |
| | $\beta$<br>(95% CIs) | P | $\beta$<br>(95% CIs) | P | $\beta$<br>(95% CIs) | P | $\beta$<br>(95% CIs) | P | $\beta$<br>(95% CIs) | P | $\beta$<br>(95% CIs) | P |
| **Autism factor**<br>**mean score\*** | 0.02<br>(-0.003, 0.04) | 0.09 | 0.003<br>(-0.02, 0.02) | 0.79 | 0.02<br>(0.003, 0.05) | 0.02 | 0.002<br>(-0.02, 0.02) | 0.88 | 0.03<br>(0.01, 0.05) | 0.004 | 0.007<br>(-0.01, 0.03) | 0.48 |

\*Standardised score, with mean = 0, standard deviation = 1 and higher scores reflecting more autism related difficulties.

#### Child’s polygenic risk for IBD, UC, Crohn’s and broad autism phenotype

There was no evidence of associations between child’s PRS for IBD, UC, Crohn’s and autism mean factor score in children (IBD: β_PRS_= 0.003; 95%CIs: -0.02 to 0.02; p= 0.79; R^2^= 0.05; UC: β_PRS_= 0.002; 95%CIs: -0.02 to 0.02; p= 0.88; R^2^= 0.05; Crohn’s: β_PRS_= 0.007; 95%CIs: -0.01 to 0.03; p= 0.48; R^2^= 0.05; Table 3, Supplementary Figure S6, Supplementary Table S13).

### Study 4: Causal effects of genetic liability to IBD on risk of autism

The mean F statistics of the IBD, UC and Crohn’s instruments were 67, 68 and 70, respectively, suggesting adequate strength. There was evidence of a causal effect of genetic liability to UC on risk of autism (_IVW_OR= 1.04; 95% CIs: 1.01 to 1.07; p= 0.006). Evidence for the effect of genetic liability to IBD and Crohn’s on autism risk was weaker, although the magnitude and direction of the effect estimates was comparable to the UC results (Table 4).

**Table 4.** Mendelian randomisation IVW estimates, 95% confidence intervals and p-values for the effect of genetic liability to inflammatory bowel disease (IBD), Crohn’s disease (Crohn’s), ulcerative colitis (UC) on autism and vice versa.

| Exposure | Outcome | OR (95% CIs) | P |
| --- | --- | --- | --- |
| Genetic liability to IBD | Autism | 1.02 (1.0, 1.05) | 0.1 |
| Genetic liability to ulcerative colitis | Autism | 1.04 (1.01, 1.07) | 0.006 |
| Genetic liability to Crohn's disease | Autism | 1.01 (1.0, 1.04) | 0.2 |
| Genetic liability to autism | IBD | 0.90 (0.73, 1.11) | 0.32 |
| Genetic liability to autism | Ulcerative colitis | 0.95 (0.77, 1.18) | 0.65 |
| Genetic liability to autism | Crohn's disease | 0.85 (0.63, 1.15) | 0.29 |

The magnitude and direction of causal effect estimates was consistent across all sensitivity analyses, and there was no evidence to suggest the influence of horizontal pleiotropy (Supplementary Table S14). Results of analyses with instruments extracted from the autism GWAS excluding ID cases were comparable to our primary effect estimates (Supplementary Table S15).

### Causal effects of genetic liability to autism on risk of IBD

The mean F statistic of the autism instruments was 28, suggesting adequate strength. There was no evidence of a causal effect of genetic liability to autism on risk of IBD, UC or Crohn’s (Table 4). The estimates were consistent across sensitivity analyses, with overlapping confidence intervals, and were unlikely to be influenced by horizontal pleiotropy (Supplementary Table S16). Repeating our analyses with instruments extracted from the autism GWAS excluding all ID cases yielded similar results (Supplementary Table S17).

## DISCUSSION

We used four complementary approaches to investigate the associations between parental diagnoses and genetic liability to IBD and offspring autism. Conducting a nationwide register-based cohort study in Sweden we found evidence of associations between parental diagnoses of IBD and offspring autism. Importantly, the maternal effect sizes were larger than paternal, without overlapping confidence intervals. PRS analyses in the ALPSAC birth cohort suggested associations between maternal genetic liability to IBD and offspring autism, while two-sample MR studies provided evidence of a potentially causal effects of genetic liability to IBD on autism risk. There was little evidence to suggest a genetic correlation between autism and IBD, as indicated by LD-score regression analyses.

The use of different study designs to triangulate our findings is a notable strength of our approach^49^. The Swedish nationwide register-based cohort study of over 2 million parent-child pairs is the largest to date on parental IBD-offspring autism. The ALSPAC cohort is a unique resource to assess polygenic associations through rich intergenerational genetic and phenotypic data. Finally, in the MR analyses we used the largest GWAS data available for all conditions and conducted several sensitivity analyses to test the robustness of our findings.

Considering study limitations, in the Swedish registers the possibility of measurement error in IBD diagnoses cannot be excluded. However, this is likely to be non-differential in relation to our study outcome and would therefore bias our findings towards the null. Secondly, while PRSs were based on large GWAS samples, it was not possible for us to investigate the variance explained by the PRSs in our target sample and the variance explained in the phenotype might be relatively small. Thirdly, in two-sample MR analyses investigating the effects of genetic liability to autism on risk of IBD, we used a relaxed instrument inclusion p-value threshold, this could potentially result in including weak instruments and therefore bias the causal effect estimates. The F statistic of the autism instruments in our analyses suggested that weak instrument bias is unlikely. Finally, using GWAS data we could only investigate the possible contribution of common variants acting under an additive model and not any contribution from rare variation which has been found to be implicated in autism^51,52^.

Larger samples and GWAS, as they become available, may help replicate and build on our findings and their potential explanations. Specifically, under liability-threshold models of inheritance, parental genetic liability to IBD could reflect sub-phenotypic manifestations of the condition^55–58^ and potentially influence fetal development in utero. These subclinical manifestations could include immune system dysregulation, micronutrient malabsorption and anaemia^,6,7,53,54^ that have been found to be implicated in IBD^5–7^ and autism^8–11^.

In conclusion, triangulating evidence from a nationwide register-based cohort study, genetic correlation, polygenic risk score analyses and MR, we found evidence suggesting associations between parental, particularly maternal, diagnoses of IBD and offspring autism. Links between maternal genetic liability to IBD and offspring autism may reflect the influence of the maternal genotype on the prenatal/intrauterine environment. Investigating the mechanisms behind these findings may provide valuable insights into the origins of autism.

## Data Availability

Data from Swedish registers and ALSPAC can be accessed after application to the respective research and ethics committees. GWAS summary data on IBD and autism are publicly available. GWAS summary data on autism excluding intellectual disabilities can be obtained after application to the iPSYCH Autism Spectrum Disorder working group.

## ACKNOWLEDGMENTS

The Medical Research Council (MRC) and the University of Bristol support the MRC Integrative Epidemiology Unit [, MC_UU_00011/1, MC_UU_00011/3, MC_UU_00011/5]. This research was funded in part, by the Wellcome Trust. For the purpose of Open Access, the author has applied a CC BY public copyright licence to any Author Accepted Manuscript version arising from this submission. CD acknowledges the support of Wellcome Trust [215379/Z/19/Z]. GDS, HJ, DR, SS, SZ are supported by the NIHR Biomedical Research Centre at University Hospitals Bristol and Weston NHS Foundation Trust and the University of Bristol. GMK acknowledges funding support from the Wellcome Trust (201486/Z/16/Z), the MQ: Transforming Mental Health (grant code: MQDS17/40), the Medical Research Council UK (grant code: MC_PC_17213 and grant code: MR/S037675/1), NIHR (project code: NIHR202646), and the BMA Foundation (J Moulton grant 2019). The iPSYCH team was supported by grants from the Lundbeck Foundation (R102-A9118, R155-2014-1724, and R248-2017-2003), NIMH (1U01MH109514-01) and the Universities and University Hospitals of Aarhus and Copenhagen. The Danish National Biobank resource was supported by the Novo Nordisk Foundation. High-performance computer capacity for handling and statistical analysis of iPSYCH data on the GenomeDK HPC facility was provided by the Center for Genomics and Personalized Medicine and the Centre for Integrative Sequencing, iSEQ, Aarhus University, Denmark. RG acknowledges funding support from the Swedish Research Council (VR2017-02900). We are extremely grateful to all the families who took part in this study, the midwives for their help in recruiting them, and the whole ALSPAC team, which includes interviewers, computer and laboratory technicians, clerical workers, research scientists, volunteers, managers, receptionists and nurses. AH was supported by grants from the South-Eastern Norway Regional Health Authority (2020022, 2018059) and the Research Council of Norway (274611, 288083).

